# Neural fingerprints of Alice in Wonderland Syndrome in patients with migraine with aura

**DOI:** 10.1101/2022.06.20.22276604

**Authors:** Giulio Mastria, Valentina Mancini, Alessandro Viganò, Claudia Piervincenzi, Nikolaos Petsas, Marta Puma, Costanza Giannì, Patrizia Pantano, Vittorio Di Piero

## Abstract

**Background:** the Alice in Wonderland syndrome (AIWS) is a transient neurological disturbance characterized by visual and somatosensory misperceptions most frequently associated with migraine. The heterogeneity of the etiologies and techniques applied to investigate the reported cases have so far prevented to draw strong conclusions about the origin of AIWS symptoms. Some lines of evidence suggest that AIWS and migraine might share common pathophysiological mechanisms, therefore we set out to investigate the common and distinct neurophysiological alterations associated with these conditions in a population of migraineurs.

**Methods:** we acquired resting-state fMRI data from 12 migraine patients with AIWS, 12 patients with migraine with typical aura (MA) and 24 age-matched healthy controls (HC). We then compared the interictal thalamic seed-to-voxel and ROI-to-ROI cortico-cortical resting-state functional connectivity between the 3 groups.

**Results:** we found a common pattern of altered thalamic connectivity in MA and AIWS, compared to HC, with more profound and diffuse alterations observed in AIWS. The ROI-to-ROI functional connectivity analysis highlighted an increased connectivity between a lateral occipital region corresponding to area V3 and the posterior part of the superior temporal sulcus (STS) in AIWS, compared to both MA and HC. The posterior STS is a multisensory integration area, while area V3 is considered the starting point of the cortical spreading depression (CSD), the neural correlate of migraine aura. This interictal hyperconnectivity might increase the probability of the CSD to directly diffuse to the posterior STS or trigger a diaschisis phenomenon causing the AIWS symptoms during the ictal phase.

**Conclusions:** taken together, these results suggest that AIWS in migraineurs might be a form of complex migraine aura, characterized by the involvement of associative and multisensory integration areas. The altered connectivity between early visual and multisensory associative areas provides a model for the pathophysiology of AIWS associated with other transient neurological conditions or with a structural etiology.

## 1. Introduction

Migraine represents the first cause of disability in young individuals and the seventh cause of disability in the global population (1,2). The wide clinical spectrum of migraine includes not only pain but also transient neurological symptoms such as visual aura that may be temporally associated with the headache phase (3). The Alice in Wonderland Syndrome (AIWS) has been recently added to this spectrum (4–7), with migraine described as the most frequent cause of this condition in adult individuals (8).

AIWS is a transient neurological disturbance causing visual and somatosensory misperceptions, with a variable combination of micro- and macropsia (seeing objects smaller or larger), telo- and pelopsia (seeing objects further or closer), macro- and micro-somatognosia (perceiving parts of the own body as bigger or smaller), and slowing in the perception of time (8–11). Since it first description by the British neurologist J. Todd (9), AIWS has been linked to several underlying etiologies (8–11), ranging from viral infections, such as Epstein-Barr (12), psychiatric comorbidities, adverse effects of common medications, stroke or brain tumors (13–15). Despite the numerous cases available in the literature (8,11), little is known about AIWS pathophysiological mechanisms.

Recently, a voxel-based metanalysis including all cases of AIWS associated with a structural lesion (16–20), showed that lesions in extrastriate visual cortex are those most frequently associated with AIWS with visual symptoms (21). To date, it remains to be elucidated if a similar brain network is implicated in AIWS caused by a non-structural disease (functional AIWS). Overall, available single-case studies in AIWS with functional neuroimaging correlates pointed to a hypoactivation of frontal regions, together with increased activation of associative parietal and occipital cortices (16–20). However, the heterogeneity of the clinical presentation, the underlying etiologies, and the variety of the methods employed to measure brain activity (i.e., fMRI, PET and SPECT) in these studies prevents from drawing strong conclusions.

Migraine might offer a homogeneous model to investigate functional brain alterations associated with AIWS, and the possibility to compare them with findings related with AIWS caused by structural brain lesions. Recent evidences highlighted a strong link between AIWS and migraine (4,22), with up to 20% of migraine patients in a tertiary referral headache clinic reporting symptoms compatible with AIWS (6). Furthermore, almost all the migraineurs with AIWS had a diagnosis of migraine with aura and a temporal concurrence of migraine attacks and AIWS episodes, suggesting that AIWS associated with migraine might be a form of complex aura with high-level cortical dysfunction (6,23).

Resting-state functional MRI (rs-fMRI) studies in patients with migraine with aura have highlighted a wide range of abnormalities in the interictal and ictal phases, including increased functional connectivity within the visual networks and other regions involved in visual processing (24,25), although this finding was not confirmed by other studies (for reviews, see Skorobogatykh et al. (26), Chong et al. (27)). The severity of these alterations has been also associated to the complexity of the migraine aura (28). Moreover, several studies have demonstrated abnormal functional connectivity (FC) between the posterior thalamus, visual cortex and precuneus in migraineurs (29–31) and more severe alterations of thalamic microstructure and structural thalamo-cortical connectivity in subjects with complex migraine aura (28).

The aim of the present study was to pinpoint the neural signatures of functional AIWS related to migraine. To do so, we compared ROI based cortico-cortical and thalamic seed-to-voxel FC of a group of patients with migraine experiencing AIWS episodes in the context of migraine attacks, a group of migraineurs with typical aura (MA) (ICHD-3 1.2.1), and a group of healthy controls (HC). We searched for common and distinct neurophysiological alterations in AIWS and MA, in order to clarify the specific mechanisms underlying AIWS in this population and their possible relationship with typical migraine aura.

## 2. Methods and Materials

### 2.2. Participants

We recruited consecutive patients with migraine with aura experiencing AIWS in the context of migraine attacks (AIWS) and sex-matched patients with migraine with typical visual and somatosensory aura (MA) at the Headache Center of Policlinico Umberto I of Rome. Because of the high within-subject aura variability (32), we included patients without a preferred side of aura symptoms.

AIWS participants and MA were recruited in the context of a cohort study at the Headache Center aimed at estimating the prevalence of AIWS in patients with migraine by means of an ad-hoc questionnaire (6). Answers to the questionnaire were validated by trained physicians and additional information about AIWS were collected, including clinical characteristics of AIWS episodes, such as the age of onset and the temporal association between AIWS, migraine and aura (6). Patients with a diagnosis of migraine with aura according to the International Classification of Headache Disorders, 3rd edition (ICHD-3), who experienced at least one episode of AIWS temporally associated with migraine attacks (i.e., from 60 min prior to pain onset to pain resolution) were included in the AIWS group. Patients with a diagnosis of migraine with aura according to the ICHD-3 without AIWS were included in the MA group. A group of sex- and age-matched healthy controls without any significant neurological or systemic disorders was blindly selected among those available from the Human Neurosciences Department archive.

The following exclusion criteria were applied to all the subjects: medically unstable or with hematological, renal, or hepatic dysfunction; history of moderate to severe head injury, stroke, or seizures; alcoholism or drug dependency.

The study was approved by the ethics committee of Policlinico Umberto I and was carried out in accordance with the latest Declaration of Helsinki. All participants provided written informed consent to the use of their data for research purpose.

### 2.3. MRI acquisition

All the images were acquired with a 3.T MAGNETOM Verio scanner (Siemens AG, Erlangen, Germany) with a 12-channel head coil designed for parallel imaging (GRAPPA) at Sapienza University of Rome. A multiplanar T1-weighted localizer image with section orientation parallel to the subcallosal line was acquired at the start of each MRI examination. Noise reduction headphones were used for attenuation of scanner noise. MRI protocol included the following sequences: 1) high-resolution 3D, T1-weighted MPRAGE sequence: TR=1900 ms; TE=2.93 ms; flip angle=9°; field of view (FOV)=260 mm; matrix=256×256; 176 sagittal slices 1 mm thick; no gap; 2) resting state functional MRI (rs-fMRI): repetition time (TR)=3000 ms; echo time (TE)=30 ms; flip angle=89°, 64×64 matrix; 50 contiguous axial slices 3 mm thick; 140 volumes (before being positioned in the scanner patients were instructed to lie down relaxed and awake with eyes closed). Dual turbo spin-echo, proton density (PD) and T2-weighted images (TR = 3,320 ms, TE1 = 10 ms, TE2 = 103 ms, FOV = 220 mm, matrix = 384 × 384, 25 axial slices 4 mm thick, 30% gap), as well as High-resolution 3D, fluid-attenuated inversion recovery (FLAIR) sequence (TR = 6,000 ms, TE = 395 ms, TI = 2100 ms, FOV = 256 mm, matrix = 256 × 256, 176 sagittal slices 1 mm thick, no gap) were also acquired to exclude concomitant brain lesions.

### 2.4. Structural images preprocessing

T1-weighted images underwent fully automated image processing with the Connectome mapper v3, a neuroimaging pipeline software combining tools such as Freesurfer v.6, FSL, ANTs, MRtrix3, Dipy and AFNI (33). Structural images preprocessing comprised skull stripping, intensity normalization, reconstruction of internal and external cortical surfaces and parcellation of subcortical brain regions (34) and further parcellation with the Lausanne atlas characterized by identical cortical regions of interest of desired size and location at different scales (from 1 corresponding to the Desikan and Killiany atlas to 5 corresponding to the smallest size (35)). The ROI-to-ROI FC analysis were conducted using the intermediate scale (scale 3, 216 cortical parcels).

### 2.5. fMRI images preprocessing

The functional images were preprocessed using SPM12 (Statistical Parametric Mapping, Wellcome Department of Imaging Neuroscience) (https://www.fil.ion.ucl.ac.uk/spm/software/spm12/). All the functional images were realigned to the first volume with a six-parameter rigid body transformation and the structural scans were then co-registered to the functional mean. The anatomical images were segmented into grey matter, white matter and cerebrospinal fluid (CSF) with the SPM12 *Segmentation* algorithm (36). The resulting images were spatially smoothed using an isotropic Gaussian kernel of 4 mm full width maximum (FWHM) and normalized to the MNI template. The choice of the smoothing parameter was based on the evidence that using smoothing kernels with a FWMH higher than 4 mm might mix the signal arising from subcortical nuclei (37).

Further steps of preprocessing and FC analyses were carried out using the CONN-fMRI Functional Connectivity toolbox v18 (https://web.conn-toolbox.org) (38). Parcellations from the Lausanne atlas based on individual anatomy were imported in *Conn* as subject-specific ROIs. Outlier scans due to head motion were identified using the software *ART* (www.nitrc.org/projects/artifact_detect) (39) and excluded if the movement of either translational or rotational parameters exceeded 2 mm or 2°. After all these quality control procedures, no subject was excluded from the following analyses.

BOLD signal noise from white matter and CSF was defined and addressed with the Component-based correction (CompCor) method, which models the influence of noise as a voxel-specific combination of multiple noise sources (38,40). The first 5 principal components of the subject-specific WM- and CSF-mask signals were calculated. Then, the linear effects of the 6 motion parameters estimated during realignment, their temporal derivatives, and the 10 noise components (including physiological artifacts such as cardiac or respiratory rates) were regressed out at each voxel. Furthermore, outliers time-points identified with the software *ART* by using a global-signal z-value threshold of 3 and a subject motion threshold of 0.5 mm were as well scrubbed from the BOLD time-series at each voxel. Finally, the BOLD time-series within each of the seeds was estimated and temporally band-pass filtered with a frequency window of 0.008 to 0.09 Hz.

### 2.6. Cortico-cortical and whole brain thalamic functional connectivity analyses

The analysis of the whole-brain thalamic FC was conducted on the images normalized in MNI space. We used a priori anatomically delineated thalami as seeds (41,42) and we merged the bilateral thalami since our patients didn’t showed a preferred side in aura and AIWS symptoms. Seed-to-voxel FC analyses were carried on between the thalami and whole brain.

For the study of the cortico-cortical FC, a ROI-to-ROI FC analysis was carried on between all the ROIs as defined by the Lausanne atlas in the native space of each subject. ROI-to-ROI FC maps were created for each participant, modeling individual-specific covariation between the BOLD activity of each ROI and that of all the other cortical ROIs by using Fisher-transformed bivariate correlation coefficients (38).

### 2.7. Statistical analyses

Second level analyses allowed to test our hypotheses and compare the FC between all the cortical ROIs in the three groups with ANOVA. False positive control in ROI-to-ROI analyses was performed using false discovery rate (FDR) correction (alpha-level = 0.05). We run separate post-hoc analyses with independent t-tests between groups for significant connections resulting from the ANOVA, correcting for the number of tests with FDR.

In the seed-to voxel analysis, we used a T-contrast to compare AIWS with HC (voxel level threshold: p < 0.001, cluster level p < 0.05 FDR corrected). We then focused on the resulting clusters of voxels and compared their FC with the thalamus between the three groups using t-tests (p < 0.05 FDR corrected).

### 2.8. Multinomial logistic regression

From both the cortico-cortical and the thalamo-cortical FC analysis we obtained a set of connections which were altered in AIWS with respect to HC and/or MA. In order to find the FC patterns which were most likely associated with AIWS, we entered the FC values of the altered connections in a multinomial logistic regression and performed model selection based on the Akaike’s information criterion (AIC) (43). We started from a full model with formula GROUP ∼C1* C2*C3 …*CN, where GROUP was a dependent categorical variable (AIWS, MA or HC), and C1 to CN, were the individual FC values, used as explanatory variables.

To test the accuracy of the resulting model we split the patients in two partitions using even and odd IDs. We used the first partition to train the model and the second to test the accuracy of model predictions. We repeated the same procedure, using the second partition to train and the first partition to test the model. Then we calculated the average model accuracy as the average of the two estimates. We further calculated the area under the curve (AUC) of the receiver operating characteristic (ROC) curve to compare model accuracy in discriminating each pair of groups.

## 3. Results

### 3.1. Participants

We recruited 12 patients with migraine experiencing AIWS (AIWS) and 12 migraine patients with typical aura (MA). Typical aura was characterized by visual symptoms (including phosphenes, photopsia and visual blurring) or visual and somatosensory (paresthesia) symptoms. A group of 24 sex- and age-matched healthy controls without any significant neurological or systemic disorders was blindly selected among those available from Human Neurosciences Department archive.

Demographic and clinical information are provided in table 1. Patients did not differ for any demographic characteristics. Characteristics of AIWS episodes are reported in table 2.

**Table 1.** Demographic and clinical information of the three groups. Mean and standard deviation and frequency counts (percentage) are reported. AIWS = Alice in Wonderland Syndrome, MA = migraine with aura, HC= healthy controls.

|  | AIWS | MA | HC |
| --- | --- | --- | --- |
| Number of patients | N=12 | N=12 | N=24 |
| Sex (female) | 75% (n = 9) | 75% (n = 8) | 70.8% (n = 17) |
| Mean age (years) | 42.9 ± 13.4 | 34.2 ± 13.2 | 38.6 ± 13.1 |
| Years of disease | 20.5 ± 11.1 | 20.8 ± 13.8 | n.a. |
| Days of migraine/month | 8.2 ± 7.3 | 4.5 ± 3.7 | n.a. |

**Table 2.** Information about AIWS characteristics in each migraineur with AIWS episodes included in this study. Age at onset refers to the age at the first episodes of AIWS. Precise age and age of onset have been replaced with an age range to prevent patients’ identification. Active AIWS indicates at least one attack of AIWS in the previous 6 months; duration of AIWS episodes refers to the average duration of the symptoms. <u>Aschematia</u> = inadequate representation of the space occupied by some parts of the body; <u>depersonalization</u> = feeling disconnected or detached from one’s body and thoughts; <u>derealization</u> = feeling dethatched from the surroundings with other people and objects seeming unreal; <u>loss of stereoscopic vision</u>: objects appearing bidimensional or flat; <u>macropsia</u> = seeing objects larger; <u>macrosomatognosia</u> = perceiving parts of the own body as bigger; <u>micropsia</u> = seeing objects smaller; <u>microsomatognosia</u> = perceiving parts of the own body as smaller; <u>pelopsia</u> = seeing objects closer; slowing in the perception of time= subjective feeling that the time is slower than expected; <u>telopsia</u> = seeing objects further. According to Lanska et al. (10) Type A = somatosensory symptoms alone; Type B = visual symptoms alone; Type C = somatosensory and visual symptoms.

| N | Sex | Age | Age at onset | Duration of AIWS episodes | Description of AIWS symptoms | AIWS type |
| --- | --- | --- | --- | --- | --- | --- |
| 1 | F | 45-49 | 25-29 | Up to 4 hours | Macrosomatognosia of face and upper limbs, mosaic vision, derealization, depersonalization, slowing in the perception of time | Somatosensory + Visual (type C) |
| 2 | F | 55-59 | 20-24 | 20-30 minutes | Micropsia, derealization, depersonalization, slowing in the perception of time | Visual (type B) |
| 3 | F | 15-19 | 15-19 | 10 minutes | Macropsia, telopsia, derealization, depersonalization, slowing in the perception of time | Visual (type B) |
| 4 | F | 45-49 | 5-9 | 15 minutes | Microsomatognosia, telopsia, pelopsia, derealization, depersonalization | Somatosensory + Visual (type C) |
| 5 | M | 25-29 | 10-14 | Up to 1 hour | Telopsia, pelopsia, mosaic vision, slowing in the perception of time | Visual (type B) |
| 6 | F | 45-49 | 15-19 | Not reported | Macrosomatognosia of hands, micropsia, macropsia, telopsia, pelopsia, derealization, depersonalization, slowing in the perception of time | Somatosensory + Visual (type C) |
| 7 | M | 35-39 | 20-24 | Not reported | Micropsia | Visual (type B) |
| 8 | F | 55-59 | 15-19 | Up to 1 hour | Micropsia, macropsia, derealization | Visual (type B) |
| 9 | F | 40-44 | 10-14 | 30 minutes | Macrosomatognosia left hemiface, macropsia, pelopsia | Somatosensory + Visual (type C) |
| 10 | F | 45-49 | 45-49 | 30 minutes | Macrosomatognosia of hands and upper limbs, aschematia, macropsia, telopsia, mosaic vision, derealization, depersonalization, slowing in the perception of time | Somatosensory + Visual (type C) |
| 11 | F | 25-29 | 10-14 | 1 hour | Aschematia, mosaic vision, derealization, depersonalization, slowing in the perception of time | Visual (type B) |
| 12 | M | 40-49 | 25-29 | Not reported | Pelopsia, mosaic vision, loss of stereotactic vision, slowing in the perception of time | Visual (type B) |

### 3.2. Whole-brain thalamic functional connectivity

We found group differences in thalamic FC in four clusters (fig.1). Thalamic nuclei were more connected with the anterior cingulate cortex (ACC: peak MNI coordinates: -2, 48, -2) in AIWS (t(19.6): 3.8, p< 0.01) and in MA (t(27.9): 2.5, p=0.017) than in HC. This hyper-connectivity was significantly higher in AIWS than in MA (AIWS vs MA: t(20.5): 5.9).

**Figure 1.**
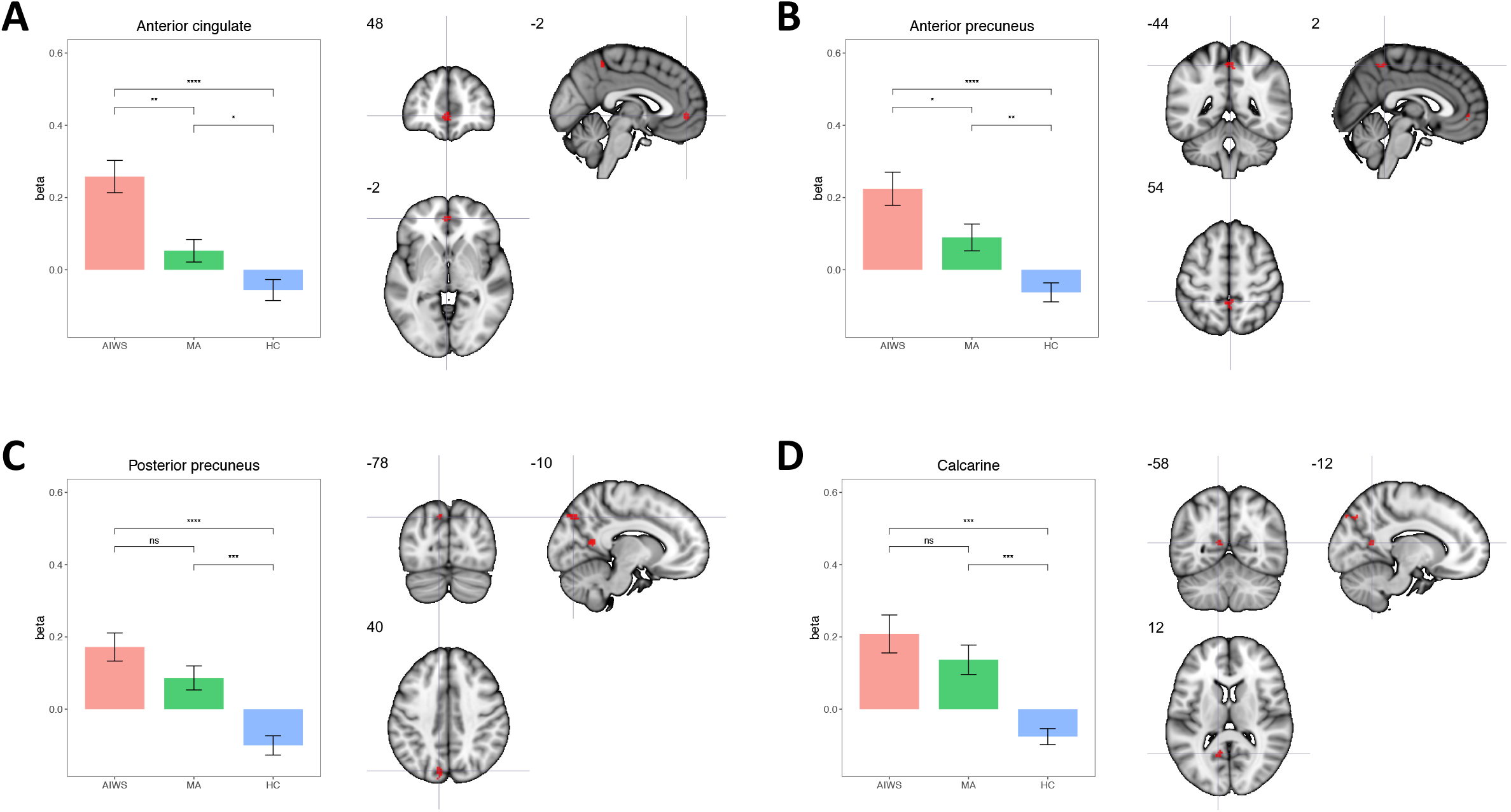
Results of the seed-to-voxel thalamo-cortical connectivity analysis. Sub-plots showing on left the mean beta values for connectivity in each group and on the right brain maps with MNI coordinates for each significant cluster: A: anterior cingulate cortex, B: anterior precuneus; C: posterior precuneus; D: calcarine.

We also found higher FC between thalamic nuclei and the anterior precuneus (AP: peak MNI coordinates: 54, -44, 2) in AIWS (t(18.4): 5.4, p<0.01) and MA (t(22): 3.3, p<0.01) than in HC, with a significantly higher FC in AIWS than in MA (t(21): 2.3, p=0.03)

The thalamic FC with the posterior precuneus (PP) and the calcarine region was higher in both AIWS (PP: t(21.4): 5.8, p< 0.01; calcarine cortex: t(20.7): 1.7, p=0.29) and MA (PP: t(21.5): 1.6, p=0.11; calcarine cortex: t(17.6): 4.6, p<0.01) compared to HC. FC of these regions, however, was comparable between AIWS and MA (PP: t(21.5): 1.6, p=0.11; calcarine cortex: t(14.9): 4.9, p< 0.01).

### 3.3. Cortico-cortical functional connectivity

At the cortical level the ANOVA found a significant group effect in the FC between a region located in the left lateral occipital cortex, corresponding to V3, and the ipsilateral posterior part of the superior temporal sulcus, which is part of the temporo-parietal junction (TPJ) (fig.2) (ANOVA: F= 19.7, p= 0.018). This difference was due to higher FC between the two regions in AIWS than HC (t(34): 5.6, p< 0.01) and MA (t(22): 4.4, p< 0.01). Conversely, HC and MA were not significantly different (HC vs MA: t(34): 0.03, p= 0.97).

**Figure 2.**
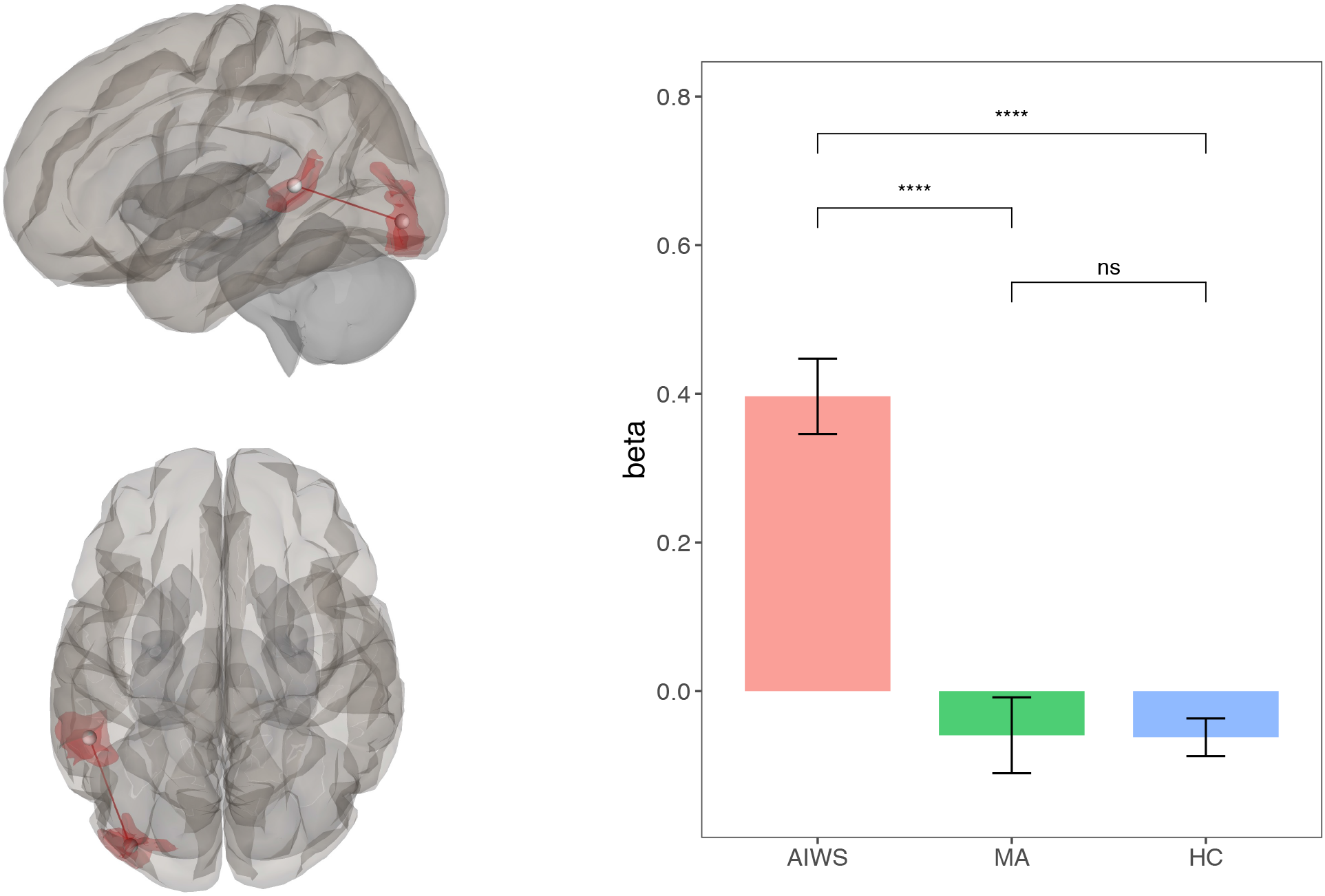
Results of the ROI-to-ROI whole brain connectivity analysis. On the right brain maps showing the statistically significant connection between left lateral occipital cortex and left superior temporal sulcus; on the left bar-plots with mean connectivity values for each group.

### 3.4. Multinomial logistic regression

In order to find the most relevant alterations characterizing AIWS with respect to HC and MA, we performed a multinomial logistic regression and performed model selection based on the AIC starting from a full model with formula: GROUP ∼C_V3-STS_* C_Thal-ACC_ *C_Thal-AP_ *C_Thal-PP_*C _Thal-Calcarine_. The final model included only three main effects (i.e., FC between V3 and STS and thalamo-cortical FC with calcarine cortex and posterior precuneus) and had formula GROUP ∼C_V3-STS_ + C_Thal-PP_ + C_Thal-Calcarine_ (Table 3). The model predicted that for a 0.1 increase in FC between the thalamus and the posterior precuneus, or between the thalamus and the calcarine cortex, the probability of being a healthy individual as compared to AIWS fell dramatically (Thal-PP: OR = 0.008, p = 0.02; Thal-Calcarine cortex: OR = 0.003, p = 0.047). Moreover, a similar increase in the FC between area V3 and STS almost doubled the risk of experiencing AIWS episodes instead of a typical aura (OR 0.476, p = 0.012).

**Table 3.** Results of the multinomial logistic regression. RRR = Relative Risk Ratio.

|  | MA |  | HC |  |
| --- | --- | --- | --- | --- |
|  | RRR | p | RRR | p |
| **Thal -**<br>**Posterior**<br>**precuneus** | 0.336 | 0.104 | 0.008 | 0.02 * |
| **Thal -**<br>**Calcarine**<br>**cortex** | 0.558 | 0.226 | 0.003 | 0.047 * |
| **V3 - STS** | 0.476 | 0.012 * | 0.576 | 0.286 |

The accuracy of the model in discriminating the patients belonging to the AIWS, MA and HC group was 83.06 %. We also calculated the AUC of the ROC curve (Figure 3, panel A), which was above 0.8 for all the pairs of groups (AUC_AIWSvsHC:_ 0.95; AUC_AIWSvsMA_: 0.86; AUC_HCvsMA_: 0.84).

**Figure 3.**
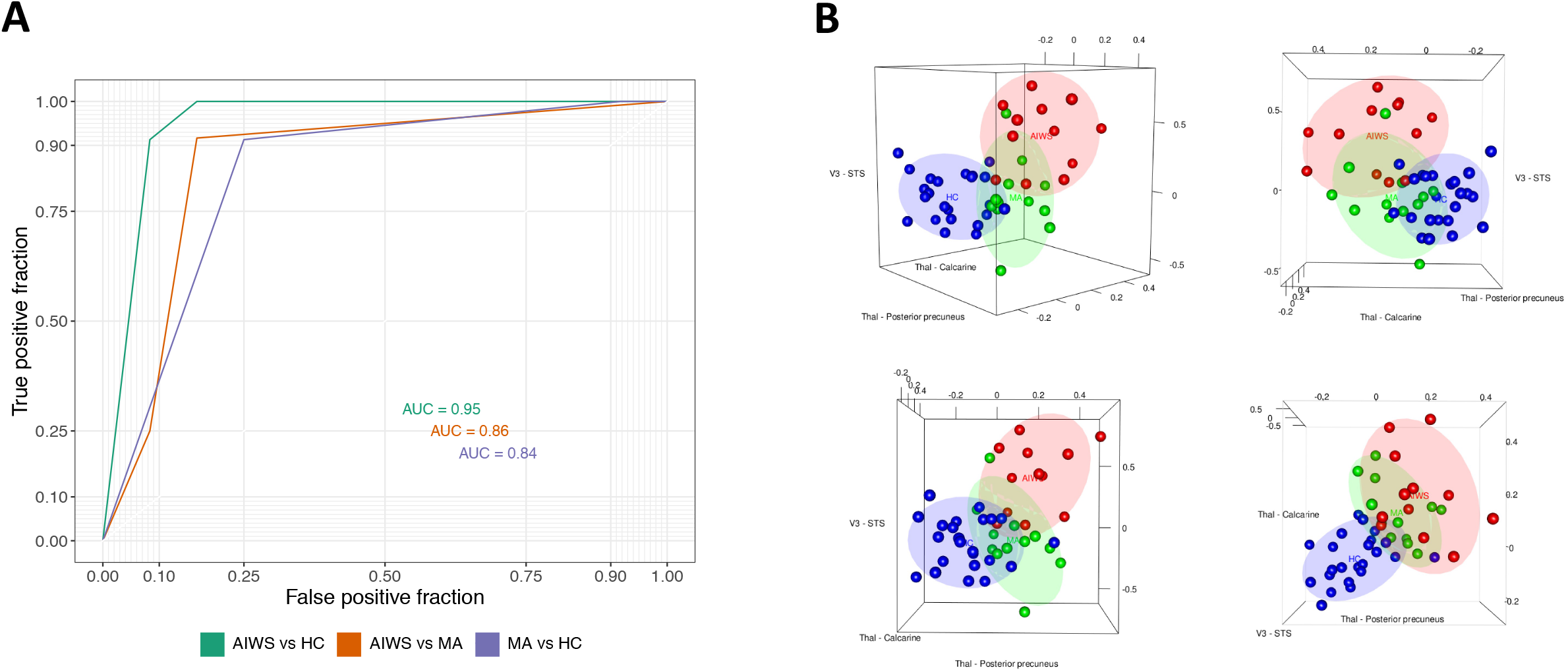
Results of the Multinomial ordinal regression and the Receive Operating Characteristic Curve (ROC) analyses. A: receiver operating characteristic curve (ROC) for prediction of AIWS diagnosis based on the connections selected with a multinomial logistic regression (i.e., V3 to STS connectivity and thalamocortical connectivity with calcarine and posterior precuneus). AUC = Area under the curve. B: The 3D scatterplots report, on each of the axis, the connectivity values of the three connections which were mostly relevant for the discrimination of AIWS (red), MA (green) and healthy control (blue). The connectivity between the thalamus and the posterior precuneus and between the thalamus and the calcarine region was more informative about the diagnosis of migraine, regardless of AIWS, with respect to healthy controls (bottom right plot). The V3-STS connectivity is more specifically related to the AIWS.

Thus, the strength of FC between the thalamus and the posterior precuneus, between the thalamus and the calcarine cortex and between V3 and STS was able to discriminate the three groups with a high level of confidence. In addition, alterations in the FC between the thalamus and posterior precuneus and calcarine regions were mostly relevant in discriminating both groups of patients with migraine from HC and did not characterize AIWS (see also Figure 3, panel B). Conversely, the hyper-connectivity between V3 and STS resulted to be distinctive of the AIWS, since it mainly contributed to discriminating between patients with migraine and AIWS from those with simple aura and healthy individuals.

## 4. Discussion

The present study provided novel evidence of interictal alterations in brain functional connectivity in patients with migraine with aura-related AIWS. We found cortico-cortical FC alterations, which are distinctive of AIWS, both compared to HC and patients with migraine with typical aura, likely representing a fingerprint of the syndrome in this population. Conversely, thalamo FC alterations seemed to be more related to migraine aura regardless of AIWS. Both AIWS and MA showed enhanced FC between the thalamus and four cortical regions, namely the anterior cingulate, anterior and posterior precuneus and calcarine cortex, with respect to HC. The increased FC between the thalamus and precuneus and calcarine regions was comparable between patients with AIWS and those with typical aura, thus likely representing a trait related to migraine with aura. The increased FC between the thalamus and anterior precuneus and ACC was more marked in patients with AIWS than in patients with typical aura, likely as a consequence of more complex visual and somatosensory symptoms in AIWS than in MA patients. In summary, none of the alterations in thalamocortical FC seems to be specifically related to AIWS, suggesting that to some extent MA and AIWS could represent a clinical continuum, and that similar pattern of FC alterations could be the common predisposing factor.

On the other hand, alterations in cortico-cortical FC appear to be specific of the AIWS. Indeed, whole brain ROI-to-ROI analysis showed an increased interictal FC between the lateral occipital cortex (V3) and the posterior STS, a part of the temporo-parietal junction (TPJ), in patients with AIWS, with respect to both MA and HC. This result may represent a functional signature of AIWS in migraineurs suggesting that the occurrence of AIWS episodes in this population, rather than other aura types, may be associated with this particular FC pattern.

Interestingly, the identified occipital region corresponds to the V3 area, which has been implicated as the origin of cortical spreading depression (CSD) (44). A recent study also showed that V3 is the central region of a wide brain network of areas characterized by volume loss in migraineurs (45), suggesting a relationship between the pattern of FC of V3 and wide-spread structural brain alterations in migraine. The posterior part of the STS is an associative area implicated in multisensory integration (46), where populations of bimodal and trimodal neurons combine auditory, visual, tactile and vestibular inputs (46,47), thus contributing to the mapping of exteroceptive stimuli in the external environment and with respect to the body (47). The involvement of the posterior STS is therefore consistent with the clinical manifestation of AIWS, which is characterized by the misperception of the distance and size of external objects and parts of one’s own body.

The AIWS patients in our sample experienced AIWS symptoms during the migraine attacks, while the present data has been collected interictally. Thus, we cannot directly infer the effect of such increased FC during the ictal phase. However, together with the temporal coincidence with the migraine attacks, the involvement of V3 points towards a possible role of the CSD in inducing AIWS symptoms. We may speculate that the interictal hyper-connectivity between V3 and STS may translate, during the ictal phase, into in an impairment of the STS through a diaschisis-like phenomenon. Alternatively, this enhanced FC may increase the likelihood of the CSD to propagate to the STS, directly impacting on its functionality.

An important limitation of our study is the relatively small sample size. This limitation is partially compensated by the fact that our groups are relatively homogeneous. Moreover, we conducted data-driven whole brain analyses applying conservative corrections for multiple comparisons. Another limitation is that we could not stratify the patients with AIWS according to their specific symptoms. Thus, we could not identify alterations specific to AIWS subtypes (10), although, at present, there is no evidence supporting the idea that different subtypes of the syndrome might be caused by different neuropathological mechanisms. Future studies with larger sample size may address this relevant question.

In conclusion, we found common and distinct alterations in functional connectivity in patients with migraine experiencing AIWS episodes and patients with typical aura during the interictal phase. We found that patients with MA and AIWS share similar alterations in thalamic FC, suggesting that these alterations are not specific of AIWS. Conversely, hyper-connectivity between V3 and the posterior STS, characterized only migraineurs with AIWS, likely representing a neural signature of AIWS in these patients. These findings are consistent with the hypothesis that AIWS might be caused by an altered FC between sensory and multisensory associative areas. Our study provides a working hypothesis for studying the pathophysiological mechanisms underlying the occurrence of AIWS episodes in patients with AIWS attributable to other etiologies than migraine, which could verify the generalizability of this hypothesis.

## Data Availability

All data produced in the present study are available upon reasonable request to the authors

